# Development of a non-infectious control for viral hemorrhagic fever PCR assays

**DOI:** 10.1101/2023.05.22.23290319

**Authors:** Matthew A Knox, Collette Bromhead, David TS Hayman

## Abstract

Assay validation is an essential component of disease surveillance but can be problematic in low resource settings where access to positive control material is limited and a safety risk for handlers. Here we describe techniques for validating the PCR based detection of *Crimean-Congo hemorrhagic fever orthonairovirus*, Ebola virus, Lassa virus, Marburg virus and *Rift Valley Fever phlebovirus*. We designed non-infectious synthetic DNA oligonucleotide sequences incorporating primer binding sites suitable for multiple assays, and a T7 promotor site which was used to transcribe the sequence. Transcribed RNA was used as template in a dilution series, extracted and amplified with RT-PCR and RT-qPCR to demonstrate successful recovery and determine limits of detection in a range of laboratory settings. Our results are adaptable to any assay requiring validation of nucleic acid extraction and/or amplification, particularly where sourcing reliable, safe material for positive controls is infeasible.

**Author Summary:** The majority of zoonoses originate in wildlife and tend to emerge from biodiverse regions in low to middle income countries, frequently among deprived populations of at-risk people with a lack of access to diagnostic capacity or surveillance. Diseases such as Crimean-Congo Hemorrhagic Fever, Rift Valley Fever, Ebola Virus Disease, Marburg Virus Disease and Lassa Fever are viral hemorrhagic fevers (VHFs) and among the most neglected and serious threats to global public health. This threat is partly due to the severity of disease caused by these pathogens, but also because their geographical distribution is close to human populations with often limited access to medical or diagnostic laboratory services. In our study we describe and validate techniques for PCR based detection of five VHF viruses using a synthetic, multi-target non-infectious positive control. Our work has applications in assay design and optimization, particularly where access to source material is problematic or requires high level biosafety containment, as is the case with VHF viruses. This approach can help learners train in techniques used in nucleic acid extraction, amplification, and sequencing of VHF viruses, but can be used for any targets, with potential for multiplexing from a single positive control.

## Introduction

Medical advances including to healthcare access and sanitation have reduced infectious disease mortality and morbidity globally. However, infectious diseases remain a significant burden to many of the most deprived people in the world, and emerging infectious diseases (EIDs) are serious public health threats, as evidenced by the emergence of SARS-CoV-2 and COVID-19 pandemic in 2019 (1). Most EIDs are zoonotic in origin, meaning they emerge from an animal and cross the species barrier to infect humans. Factors that lead to EID emergence include socioeconomic factors, land use change, and urban population growth (2-5). The majority of zoonoses (e.g. ∼72% (3)) originate in wildlife and because of these processes they tend to emerge from biodiverse regions in low to middle income countries (LMICs) and frequently among deprived populations of at-risk people with a lack of access to diagnostic capacity or surveillance. The recent global emergence of monkeypox virus highlights the diagnostic issue further (6); the virus that causes Mpox (monkeypox), an endemic zoonosis in West and Central Africa, must have emerged locally, possibly in Nigeria, and gone undetected until it was detected in Europe (6).

Diseases such as Crimean-Congo Hemorrhagic Fever (CCHF), Rift Valley Fever (RVF), Ebola Virus Disease (EVD), Marburg Virus Disease (MVD) and Lassa Fever (LF) are viral hemorrhagic fevers (VHFs) and among the most neglected and serious threats to global public health. This is in part due to the severity of disease caused by these pathogens, but also because their geographical distribution is close to human populations with often limited access to medical or diagnostic laboratory services. The large West African EVD outbreak was likely able to establish during the months after the first case because early cases were not detected in areas poorly served by diagnostic services (7). There is therefore an urgent need to develop tools for surveillance and diagnosis for both people and wildlife hosts for the VHFs (8).

Numerous diagnostic assays exist for the viral agents of these VHFs (9-14). However, there are difficulties in training personnel, particularly for VHFs and pathogens that are both rare and highly pathogenic (15-17). Developments in synthesizing nucleic acids, however, allow the synthesis of non-infectious control material for safe handling, including in low resource settings (18, 19). Here, we designed synthetic DNA oligonucleotide sequences incorporating primer binding sites suitable for five VHF (CCHF, RVF, EVD, MVD, LF) causing viruses including a T7 promotor site to transcribe the sequence to enable RNA synthesis and provide positive control material for all laboratory steps from nucleic acid extraction to detection. We tested the sensitivity of extraction using both RT-PCR and RT-qPCR. In addition, since access to refrigeration and cold storage is often not possible in field settings, we assess the stability of reagents for amplification after storage at ambient temperature over a one-week time course.

## Methods

### VHF Control Vector

A 1,000 bp DNA fragment was designed by generating random nucleotide sequence and embedding previously published primer binding sites (9-14) within the sequence at positions designed to amplify the target amplicon size for each assay. The fragment contained primer binding positions for the amplification of five negative strand RNA viral pathogens, comprising three viruses from the order Bunyavirales: *Crimean-Congo hemorrhagic fever orthonairovirus* (CCHFV, family Nairoviridae), Lassa virus (LV, *Lassa mammarenavirus*, family Arenaviridae), and *Rift Valley fever phlebovirus* (RVFV, family *Phenuiviridae*), and two viruses from the order Mononegavirales: Ebola virus (EV, *Zaire ebolavirus*, family Filoviridae) and Marburg virus (MV, *Marburg marburgvirus*, family Filoviridae) (Figure 1, Table 1).

**Figure 1.**
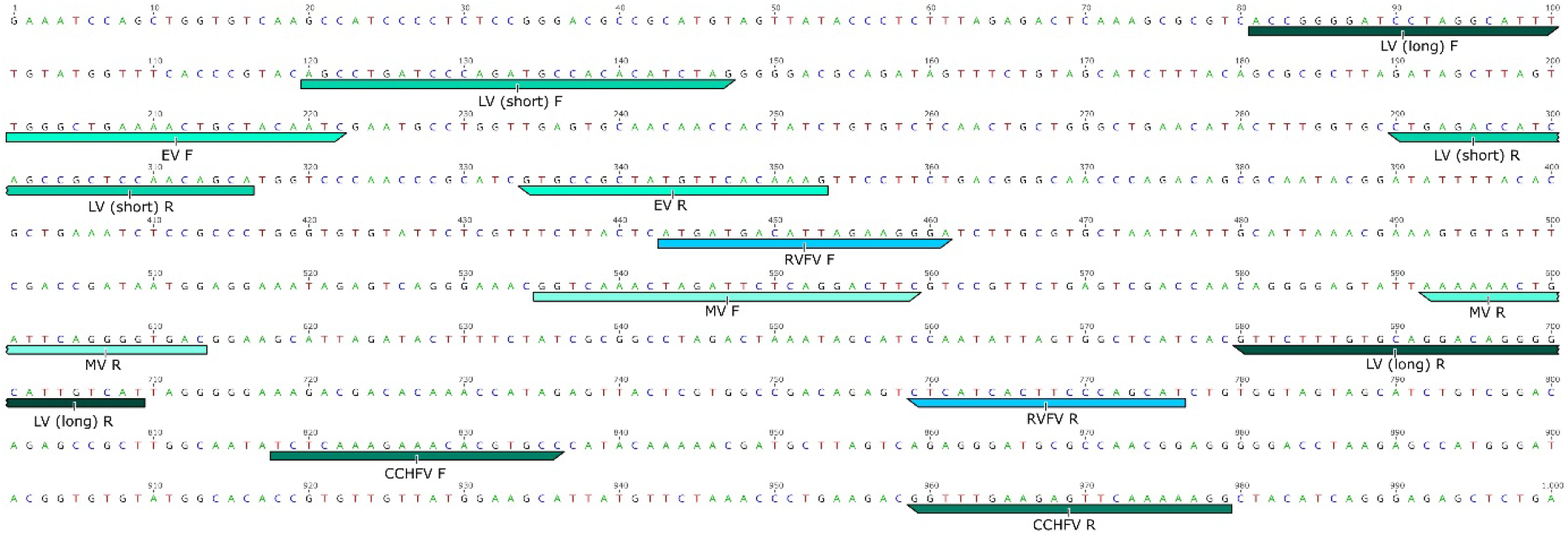
Viral hemorrhagic fever synthetic insert with primer sequence locations. See text and Table 1 for details.

**Table 1.**
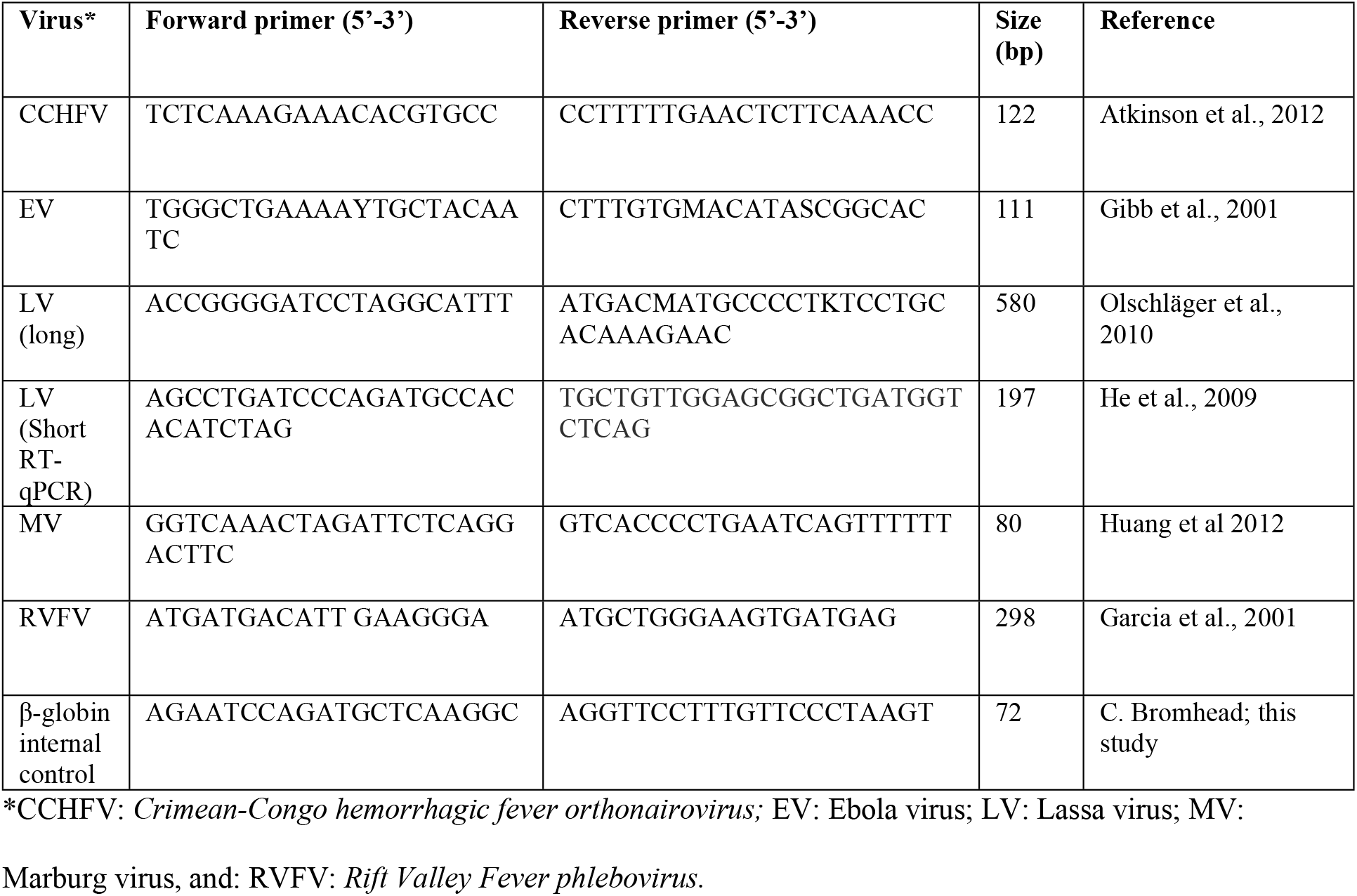
Primers and probes for RT-PCR and RT-qPCR

The synthesised product was manufactured by GeneScript and cloned using pET-20b(+) vector, which includes a T7 promoter site (Figure 2). The resulting lyophilized plasmid DNA was reconstituted in 20 μL sterile water. RNA copies of the fragment were generated from plasmid DNA using the MAXIscript T7 In Vitro Transcription kit (Ambion) following the manufacturer’s instructions. Synthesized RNA was then quantified using Qubit™ RNA HS (High Sensitivity) Assay Kit and stored at -80°C until further analyses. We estimated the copy number based on the calculation which can be found at http://www.scienceprimer.com/copy-number-calculator-for-realtime-pcr.

**Figure 2.**
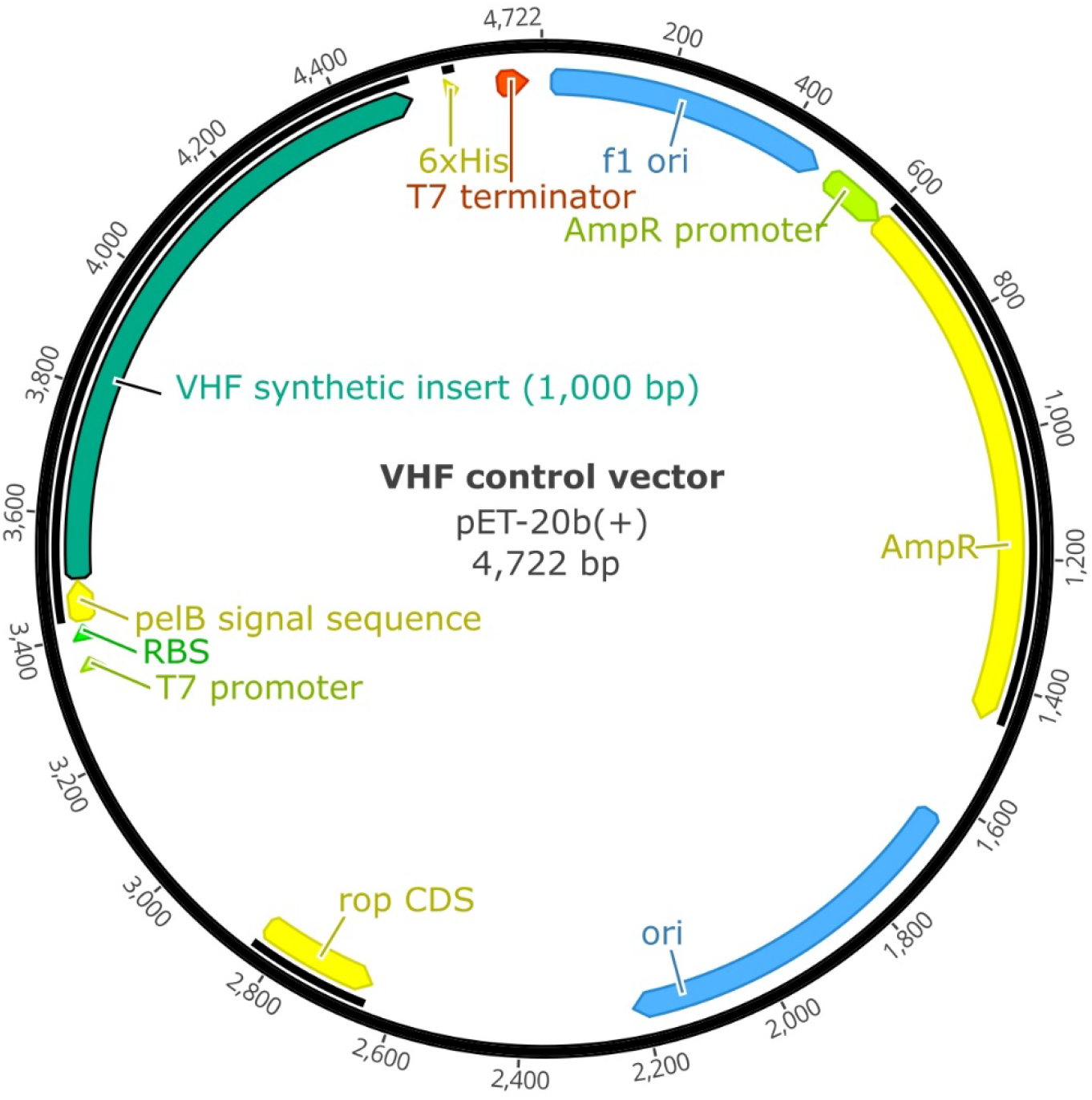
pET-20b(+) vector including insert, which is the sequence in Figure 1.

### RNA extraction and 1-step RT-PCR

To simulate the biological sample processing steps, the synthetic VHF control RNA (550 - 5.5×10^11^ copies) was added to 1 mL sterile PBS and then passed through a sterile 0.45 μm filter (Macherey-Nagel GmbH & Co. KG, Düren, Germany). We used 200 μL of filtrate as the input material for nucleic acid extraction using the High Pure Viral Nucleic Acid kit (Roche, New Zealand) according to the manufacturer’s procedure. Extracted RNA was amplified by 1 step RT-PCR using each of the five VHF primer sets (Table 1) and assay-specific cycling protocols (Table 2). Each 20 μL reaction consisted of 0.25 μM each primer, 1 pg-1μg of template RNA (VHF Control Vector, GeneScript USA), 1 x PCR buffer and 0.5 μL SuperScript™ III RT/Platinum™ Taq High Fidelity Enzyme Mix (Invitrogen). RT-PCR products were separated by agarose gel electrophoresis and visualised under UV light where bands of the expected size were identified and excised. DNA was eluted in 50 μL buffer (10 mM Tris, pH 8.0) for 12–24 hours at 4°C and then sent for bi-directional Sanger sequencing to the Massey Genome Service (Massey University, Palmerston North, New Zealand).

**Table 2.**
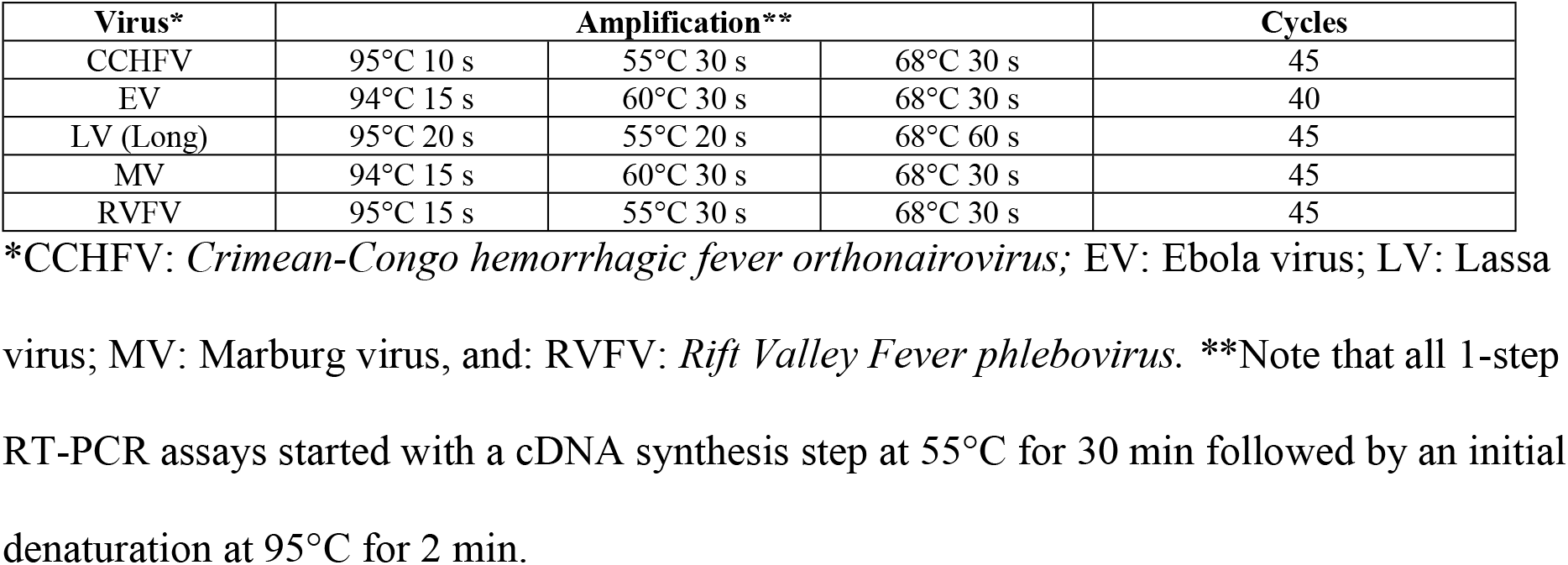
Cycling conditions for 1-step RT-PCR assays

To test the sensitivity of the nucleic acid extraction and 1-step RT-PCR assays, we conducted a dilution series with the synthesized RNA. Dilutions for extraction were prepared in PCR grade water and ranged from 31.2 to 3.12×10^−8^ ng (copy numbers 5.5×10^11^ to 550) and followed the same biological sample processing steps as above.

### RT-qPCR assay and reagent stability experiments

All RT-qPCR assays used Ultraplex 1-Step ToughMix (Quantabio) and Eva®Green 20x (Biotium, both from DNature NZ Ltd) on a Roche LightCycler-96 instrument. We included two additional primer sets for RT-qPCR assays: a shorter Lassa virus amplicon (14) and a beta-globin internal cellularity control (β-globin) which we found could be duplexed with each VHF assay. Cycling conditions were optimised by testing annealing temperature gradients from 55°C to 60°C in duplicate for all seven assays under the following thermocycler conditions: 50°C for 10 mins, 95°C for 3 mins, 55 cycles of 95°C for 5 s, 55-60°C for 15 s, 72°C for 30 s, with fluorescence acquired during the extension step (excitation/emission Eva green = 488/530 nm). See Table 3 for optimal run conditions for each VHF. Primer concentrations were optimised by titration from 0.2 to 0.6 μM in 0.05 μM increments using a dilution series from 5000 copies/μL to 0.5 copies/μL of the synthetic control extracted as for test samples (High Pure Viral kit, Roche New Zealand). The dilution series was used to determine the limit of detection of each assay (see results, Table 3). Optimal reagent conditions for each VHF+ β-globin RT-qPCR assay, in a 10 μL reaction, consisted of 1 x Toughmix 1-step buffer, 1 x Eva green dye, 0.45 μM each primer (VHF + β-globin), synthetic RNA control (50-5000 copies/μL) and RNAse-free water.

**Table 3.** RT-qPCR VHF virus assay optimal experimental conditions and limits of detection.

| Target* | Optimal annealing temperatures ($^{\circ}\text{C}$) | TM of melting peaks ($^{\circ}\text{C}$) | Limit of detection (copies/$\mu\text{L}$) |
| --- | --- | --- | --- |
| EV | 55-61 | 82.5 | 50 |
| RVFV | 55-56 | 82.5 | 50 |
| CCHFV | 55-62 | 83 | 50 |
| MV | 55-61 | 78 | 50 |
| LV-long | 55-65 | 77 | 500 |
| LV-short | 55-58 | 81 | 50 |
| β-globin | 55-60 | 81 | 10 |

\*CCHFV: EV: Ebola virus; RVFV: *Rift Valley Fever phlebovirus*; *Crimean-Congo hemorrhagic fever orthonairovirus*; MV: Marburg virus, and: LV: Lassa virus.

To assess the performance of our assays under potential field conditions where refrigeration may not be available, we tested the performance of RT-qPCR reagents Ultraplex 1-Step ToughMix and Eva®Green 20x stored at room temperature (12-28°C) for up to 1 week before use. Duplicate RT-qPCR tests for each VHF were performed at each of three timepoints (24 hours, 72 hours and 168 hours, 7 days) using the room temperature reagents. Synthesised RNA from the VHF Control Vector was used as the template at a concentration of 1,600-2,500 copies/μL.

## Results

### Dilution series RNA extraction and RT-PCR

RT-PCR performance varied among the assays (Supplementary Table 1, Supplementary Figure 1). The results of a dilution series of synthetic control from 5.5E+11 to 552 copies showed all five assays could detect to a limit of ∼5.5 million copies/μL by gel electrophoresis, while the LV and MV could reliably detect 5.5E+4 and gave visible weak bands at ∼550 copies/μL. Sequencing of excised amplified products confirmed that recovered DNA matched target amplicons (data not shown).

### VHF RT-qPCR assay and time course experiment

The RT-qPCR assays underwent optimisation for annealing temperatures, primer concentrations and high-resolution melt (HRM) analysis. The results for these parameters as well as the limits of detection for each RT-qPCR assay is shown in Table 3 and Figure 3.

**Figure 3.**
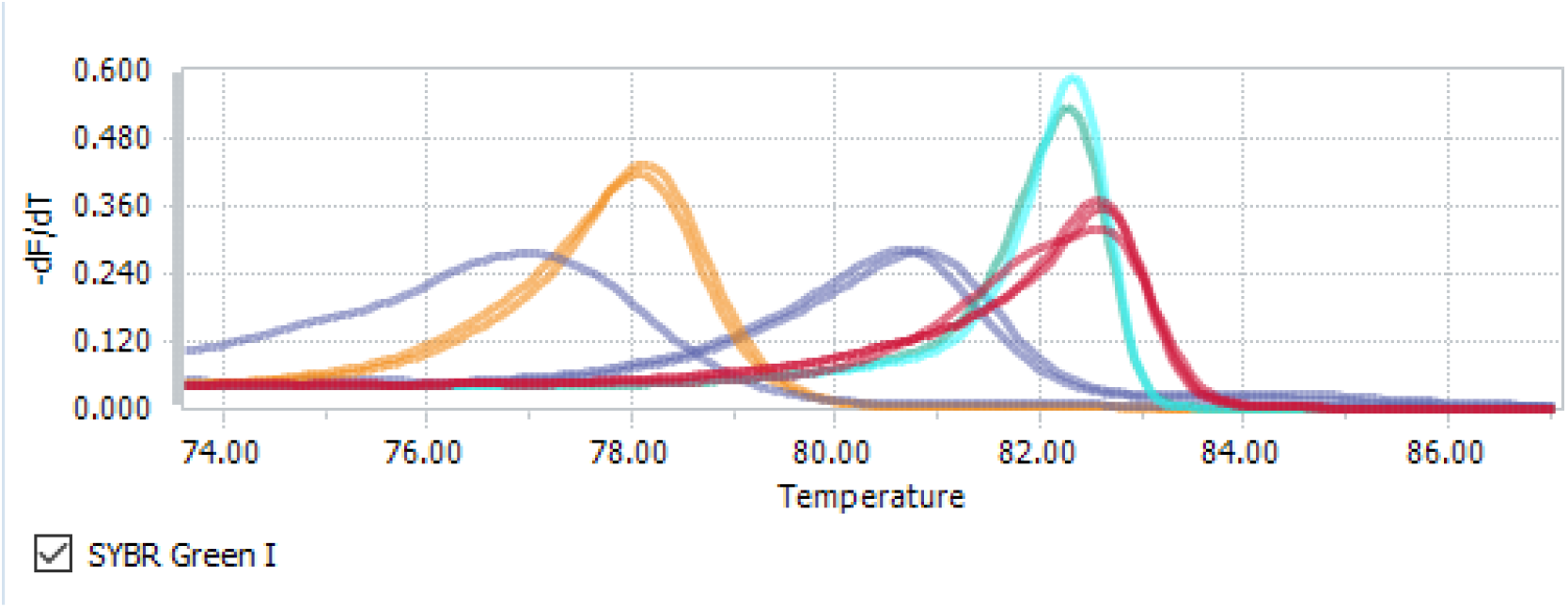
High resolution melting peaks for EV and CCHFV (red), RVFV (turquoise), MV (yellow) Short LV and β-globin (81°C purple), Long LV (77°C purple). Colours are grouped by melting temperature (°C).

All assays could reliably detect 50 copies per μL except for the long LV assay which could only detect down to 500 copies/μL reliably. The decreased efficiency of this assay indicated it was not suitable for qPCR conversion. No change in primer concentration (0.45 mM) was required for combining a VHF assay with the β-globin internal control (data not shown) and a standard annealing temperature of 56°C was found to suit all assays.

We tested the performance of our VHF assays using RT-qPCR reagents stored at room temperature (12-28°C), in the dark, for up to 1 week. Duplicate RT-qPCR tests for each VHF were performed at each of three timepoints (24 hours, 72 hours and 7 days) using the synthetic RNA control at a concentration of 1,600-2,500 copies/μL.

The average Cq values at 24 hours, 72 hours and 1 week all rose compared to time zero (using ideal frozen reagents). The VHF virus RT-qPCR’s biggest change was between 72 hours and 1 week for each VHF virus assay (Figure 4). The average Cq values were highest for the LV assays, particularly the Long LV, indicating it was the most sensitive to reduced efficiency reagents. By contrast, the MV assay maintained a low Cq across all three time points. After 1 week, assay sensitivity for detecting 1000 copies/μL is significantly compromised. Therefore 3 days (72 hours) at room temperature is the maximum recommended storage time for Ultraplex 1-Step ToughMix (Quantabio) and Eva®Green 20x (Biotium) for this testing purpose (See Supplementary Table 2 for the raw Cq data).

**Figure 4.**
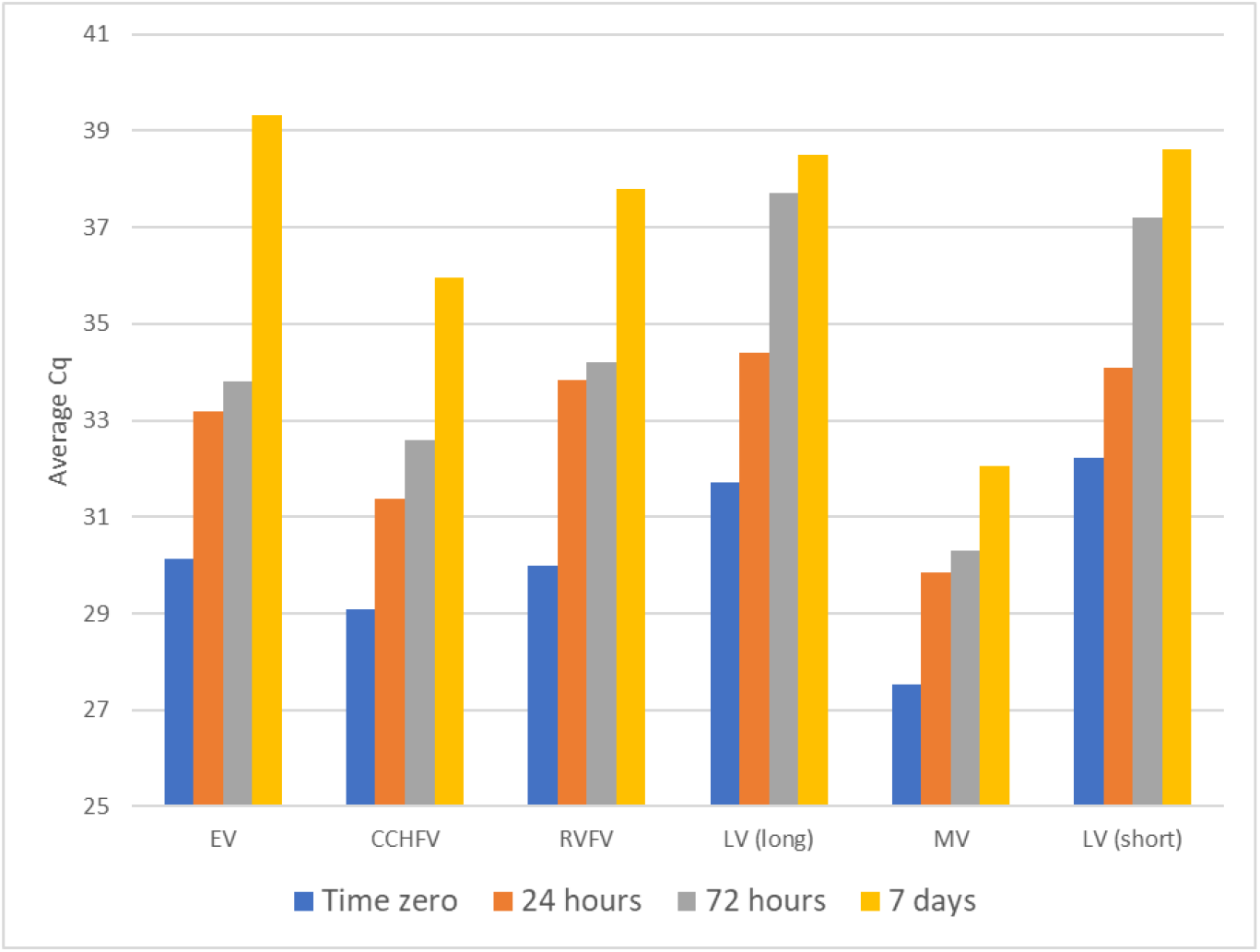
Room temperature stability of ToughMix and Eva Green for VHF virus RT-qPCR assays.

## Discussion

We have developed six assays for the most commonly occurring viral causes of VHF’s using both RT-PCR and RT-qPCR, incorporating an internal control for the latter, with the possibility of multiplexing and using reagents stored at room temperature for up to 72 hours. Our results demonstrate the applicability of synthetically designed nucleic acids for use as molecular diagnostic assay controls in both RT-PCR and RT-qPCR methods. This work has applications for assay design and optimization, particularly where access to source material is problematic or requires high level biosafety containment, as is the case with VHF viruses.

This approach can help learners to train in techniques used in nucleic acid extraction, amplification, and sequencing of VHF viruses, however, as well as providing ongoing validation for the VHF viral assays we present, this approach can be used for any targets, with potential for multiplexing from a single positive control.

Recent technological advances have greatly improved virus detection and diagnosis without the need for multi-step RNA purification (20). New generation RT-qPCR reagents are more robust to temperature storage above -20°C. These tests provide rapid, inexpensive, and robust diagnoses where laboratory infrastructure is not available, and may replace current technologies (i.e., RT-PCR assays) in some settings. However, there is an urgent need to develop in-country tools for surveillance and diagnosis for both people and wildlife hosts for the VHFs (8). Since our workflow requires only standard equipment, currently present in most molecular biology labs it has applications for the design and validation of tests used in surveillance of VHFs in countries where such work is most needed and for the time being RT-PCR is likely to remain the most common method for many diagnostic laboratories. Therefore, safe approaches to generate control material and to train staff are needed and our work provides further evidence for the applicability of synthetic nucleic acids for use as assay controls.

## Data Availability

All data used in the submission can be accessed in the tables, figures and supplementary data provided in the submission.

## Acknowledgements

This work was supported by funds from the World Organisation for Animal Health (Grant 3000034275; OIE Laboratory (or Collaborating Centre) Twinning Project: Enhancing capacity for early detection of viral haemorrhagic fevers in Liberia through epidemiological and laboratory training), Royal Society Te Apārangi Rutherford Discovery Fellowship (RDF-MAU1701) and the Percival Carmine Chair in Epidemiology and Public Health (DTSH). We wish to acknowledge the valuable contributions of WOAH staff, particularly Sophie Muset and Mariana Marrana, and the Central Veterinary Laboratory team in Liberia.

## CreDit Author Contributions

**Matthew A Knox:** Investigation, Methodology, Formal Analysis, Visualization, Writing – Original Draft Preparation.

**Collette Bromhead:** Investigation, Methodology, Validation, Formal Analysis, Writing – Review & Editing.

**David TS Hayman**: Conceptualization, Funding Acquisition, Project Administration, Writing – Review & Editing.

